# High-intensity focused ultrasound in treatment of primary breast cancer: a systematic review and meta-analysis

**DOI:** 10.1101/2024.09.10.24313423

**Authors:** Sogol Alikarami, Hamid Harandi, Ali Jahanshahi, Seyed Sina Zakavi, Negin Frounchi, Mehrdad Mahalleh, Sara Momtazmanesh

**Affiliations:** School of Medicine, Tehran University of Medical Sciences, Tehran, Iran; School of Medicine, Guilan University of Medical Sciences, Rasht, Iran; Liver and Gastrointestinal Disease Research Center, Tabriz University of Medical Sciences, Tabriz, Iran; Kidney Research Center, Tabriz University of Medical Sciences, Tabriz, Iran; Department of Musculoskeletal Radiology, Russell H. Morgan Department of Radiology and Radiological Science, Johns Hopkins University School of Medicine, Baltimore, MD, USA

**Keywords:** High-Intensity Focused Ultrasound Ablation, Non-invasive therapies, breast cancer, focused ultrasound surgery

## Abstract

**Background:** In recent years, the tumor management strategies have focused on less invasive methods, aiming to yield optimal efficacy while minimizing further complications and enhancing the overall outcome of patients. High-intensity focused ultrasound (HIFU), a known thermal ablative technique, has shown promising results in breast cancer treatment. Therefore, we performed this systematic review and meta-analysis to assess the clinical, histopathologic, immunologic, and radiologic outcomes of HIFU ablative therapy and its complications in patients with primary breast cancer.

**Methods:** We searched PubMed and Scopus databases to identify the eligible articles. Data extraction was conducted by two independent authors. A random effect model was employed to pool the proportion of remaining tumor after HIFU therapy in breast cancer. Pooled CD4/CD8 ratio mean difference between HIFU and radical mastectomy was, measured using a fixed-effect model.

**Results:** We included 26 studies and 677 participants in the systematic review. Tumor necrosis rates varied, with 4 studies reporting less than 50% complete necrosis and 5 more than 50%. Two studies observed HIFU-induced disturbances in microvasculature of the targeted tissue. Six noted no contrast enhancement in successfully treated areas, two observed a thin rim indicating necrosis or fibrosis, and four reported a persistent enhancement in MRI images associated with a residual viable tumor. The weighted proportion of patients with residual tumor was 58.45 (95% C: 45.48 – 71.42). The CD4/CD8 ratio was higher in the HIFU group, with a weighted mean difference of 0.6 (95% CI: 0.41 – 0.78). The most prevalent side effects were pain (47.14%) and skin burn (2.59%).

**Conclusions:** HIFU is a relatively safe procedure for treatment of breast cancer as an independent or conjugated therapy and its effectiveness is promising regarding histopathological response, immunological reactivity, and vascular damage in the targeted area.

## 1. Introduction

Breast cancer is the most common type of malignancy worldwide. In 2015, 623,000 deaths were reported due to breast cancer, making it the fifth leading cause of cancer-related deaths. Between 1990 and 2017, the incidence of breast cancer increased by 123% (1). In 1882, Halsted proposed an innovative surgical method for breast cancer treatment. This technique, known as radical mastectomy, includes a unilateral axillary complete resection of breast tissue with overlying skin, pectoral muscles, and ipsilateral axillary lymph nodes (2). In 1948, Patey and Dyson introduced modified radical mastectomy as an alternative to Halsted’s, consisting of total mastectomy with total axillary dissection and preserved pectoralis major muscle, making it less invasive (2).

In 2002, a 20-year follow-up of the National Surgical Adjuvant Breast and Bowel Project (NSABP) clinical trials revealed no significant difference in survival rates for mastectomy in comparison to lumpectomy with or without postoperative breast irradiation. However, after lumpectomy, breast irradiation significantly decreased the recurrence in the ipsilateral breast (3). This finding was a fundamental step towards locoregional, non-operative, and less invasive breast cancer treatment techniques and breast conservation as an indicator of treatment quality (4). Alternatives to invasive methods need to provide more accurate margins, intraoperative tumor imaging capability (5), fewer complications from surgery and general anesthesia, faster postoperative recovery, lower re-excision rates, and better cosmetic outcomes while maintaining efficacy and enhancing the quality of life after the treatment (6).

Thermal ablative techniques, i.e., cryoablation, laser ablation, microwave ablation, radiofrequency ablation, and high-intensity focused ultrasound (HIFU), induce necrosis of a target lesion using extreme hyperthermia or hypothermia (7). HIFU, also called focused ultrasound surgery (FUS), is an entirely non-invasive technique progressively used not only for benign conditions such as uterine fibroids and breast fibroadenomas(8) but also for the treatment of the pancreas, bone, connective tissue, liver, thyroid, parathyroid, kidney, and brain malignancies as an alternative or in combination with already established treatments (9). In this modality, a piezoelectric ultrasound transducer propagates a high-amplitude pressure wave, increasing targeted tissue temperature above 55 °C, resulting in heat coagulation (10). This technique is combined with ultrasound or magnetic resonance imaging (MRI) as guidance for precise targeting of the tissue and evaluating outcomes (6).

In this systematic review, we intend to assess immunologic and histopathologic responses to HIFU ablative therapy in the treatment of primary breast cancer and evaluate its outcomes based on clinical and radiological perspectives. Furthermore, in the current study, we will address the side effects of this method.

## 2. Material and Methods

This systematic review was performed consistent with Preferred Reporting Items for Systematic Reviews and Meta-Analyses (PRISMA) guidelines (11). Moreover, the review protocol was registered in the International Prospective Register of Systematic Reviews (PROSPERO) (https://www.crd.york.ac.uk/prospero/display_record.php?ID=CRD42024499582).

### 2.1. Search strategy and study selection

A systematic search was conducted in the PubMed and Scopus databases on December 23, 2023 using keywords related to ‘breast neoplasms’ and ‘high-intensity focused ultrasound ablation’ along with their MeSH terms. The details of the search entries can be observed in Supplementary table 1. We also manually searched other resources, such as websites, organizations, and citation list of related articles. After removing duplicate results, two reviewers independently screened articles based on the title and abstract, and then the full texts were assessed. Any conflicts or discrepancies were resolved through discussion or a third investigator during the search process.

### 2.2. Inclusion and exclusion criteria

All original articles containing information about HIFU ablative technique outcomes and complications in patients with primary breast cancer until December 23th, 2023, were included, except case reports and case series with less than 5 participants. Non-clinical and non-human studies, studies about benign lesions, review articles, conference papers, letters to editors, study protocols, and non-English literature were excluded.

### 2.3. Data items

Two independent reviewers extracted the following data from the included studies. The studies and patients’ characteristics consist of the first author’s name, year of publication, country, number of participants, and their mean age, tumor size, and comparator. Device model and producer, power, guidance modality, ablation margin, and time were extracted as HIFU-based treatment characteristics. Any disagreements were resolved by a third reviewer through discussion.

### 2.4. Quality and publication bias assessment

The quality of the included studies was assessed by two reviewers separately using the ROBINS-I score for nonrandomized studies. This scoring system consists of three domains: pre-intervention, intervention, and post-intervention. Accordingly, the studies were categorized as low, moderate, serious, or critical risk of bias or no information (12). In randomized control trials, the risk of bias was estimated by the RoB 2 tool made up of five components. All randomized articles were classified as low risk of bias, some concerns, or high risk of bias (13). Any conflicts were solved through discussion with a third reviewer. To reduce the impact of publication bias, we examined the reference lists of the studies included in our analysis.

### 2.5. Statistical analysis

All collected data were organized into tables and presented as means ± standard deviations (SD), medians with ranges or interquartile ranges, and percentages. We performed a single-arm proportional meta-analysis to measure the pooled percentages of patients with residual tumor after performing the HIFU after at least 1 years of follow-up. Furthermore, we implemented a fixed-effect model to compare the mean differences of CD4/CD8 ratios between HIFU groups and radical mastectomy participants. All statistical analysis were preformed using Stata version 17.0 software (StataCorp LP, College Station, Texas, USA). P value of <0.05 was defined as statistical significance.

## 3. Results

### 3.1. Study selection

We achieved a total number of 1417 records based on our systematic search. After removing duplicates, 960 articles underwent the title/abstract screening process, of which 113 reports were found eligible for detailed evaluation. From these, we excluded 88 articles due to the following reasons: non-clinical studies (n=29), irrelevancy (n=22), assessing benign lesions (n=13), non-English records (n=2), review articles (n=13), conference papers (n=4), case studies (n=2), letters (n=1), and study protocols (n=1). Another study was excluded because it reviewed the two included studies and reported their data (15). Furthermore, one study was identified through citation searching. At last, 26 studies (16–41) were included in our systematic review based on our eligibility criteria. One study by Wu and colleagues (29) had the same population, with another included study (30) reporting different data from them. Therefore, we included both studies and considered the participants as one study (Figure 1).

**Fig 1.**
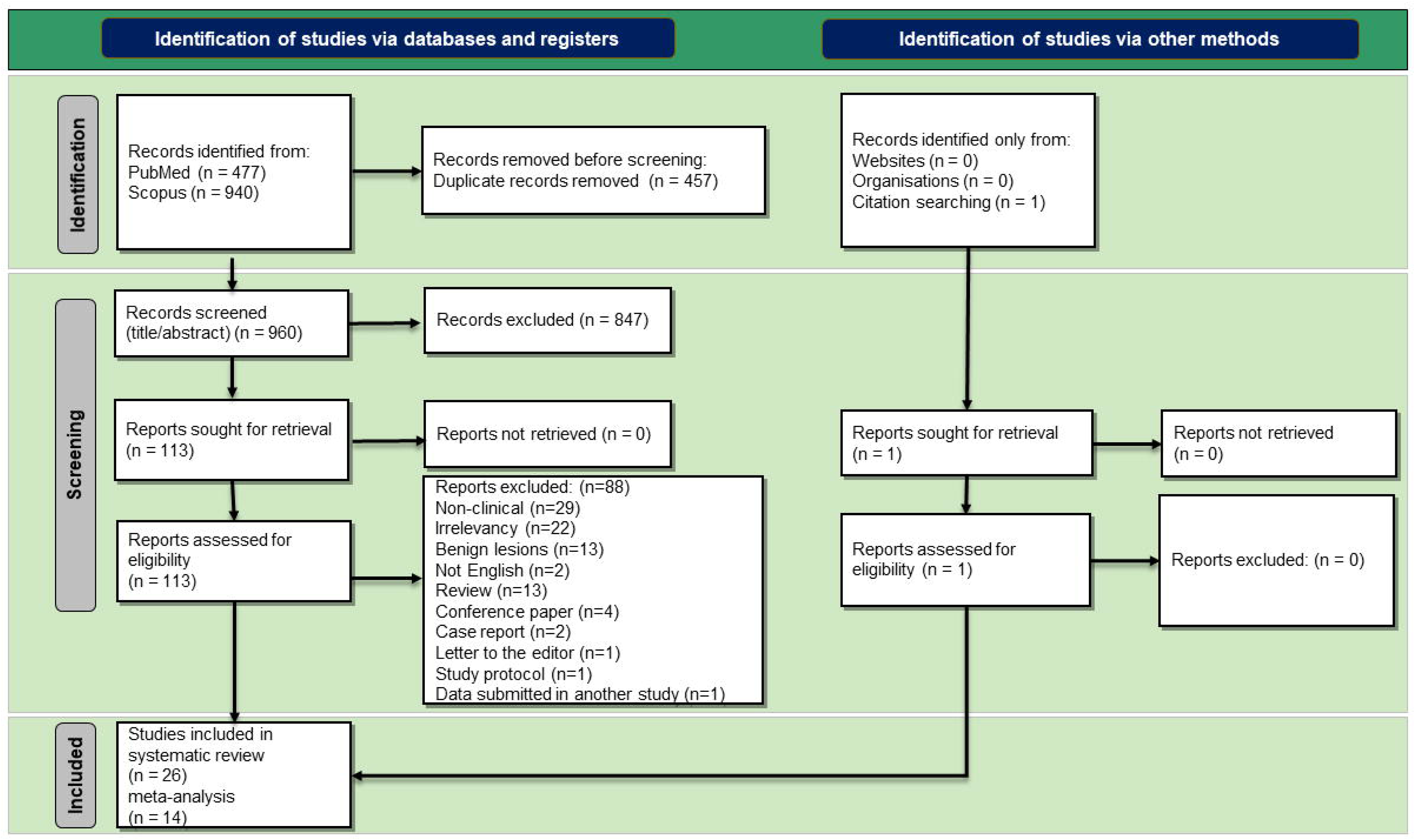
PRISMA Flow Diagram for Study Selection Process

### 3.2. Overview of the studies’ characteristics

Of the 26 included studies, 14 were from China (16,17,21–24,27,29–32,35,38,40). Thirteen studies were clinical trials (21–25,29,31,32,34,35,38–40). The total number of participants was 677, of which 479 were for the intervention groups. The number of participants who underwent HIFU in each study varied from 6 (33) to 50 (35), with a mean age ranging from 44.7 (35) to 74.2 (20) years. Ten studies had a comparator group in their study (21–23,31,32,34–36,38,40), from which seven studies (21– 23,31,32,38,40) used patients who underwent modified radical mastectomy (MRM) as their control group. However, the comparator of one study was radiofrequency ablation (36), another considered neoadjuvant chemotherapy (35), and Deckers et al. (34) used healthy female volunteers as their control group. Only nine studies (18,19,26,28,31,37,38,40,41) specified the breast cancer type, which included infiltrating ductal carcinoma, infiltrating lobular carcinoma, invasive and non-invasive ductal carcinoma, invasive lobular carcinoma, ductal carcinoma in situ, lobular carcinoma in situ, medullary carcinoma, and mucinous carcinoma. Twenty studies (16,17,19–25,27–33,35,38–40) reported their ablation margin diameter, which ranged from 0.1 cm (20) to 2.2 cm (23). Fourteen studies reported an ablation margin between 1-2 cm (16,17,21,22,24,27,29–33,35,38,40). However, two studies (19,28) mentioned their ablation margin was 0.5 cm. Gianfelice et al. reported their mean ablation margin was 0.35 cm (20), while Payne and colleagues and Zippel et al. stated that their margin for ablation was 0.4 cm and at least 0.5 cm, respectively (25,39). Eleven studies (18–20,25,26,28,34,36,37,39,41) implemented MRI as their guidance modality for HIFU ablation, while others performed an ultrasound-guided procedure. More detailed information about the studies’ characteristics is illustrated in Table 1.

**Table 1.**
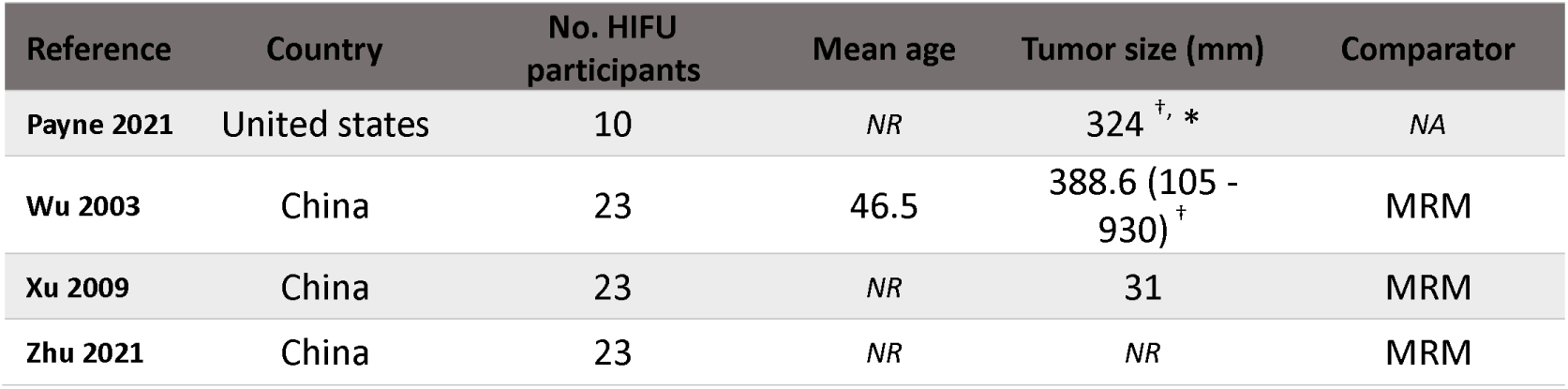
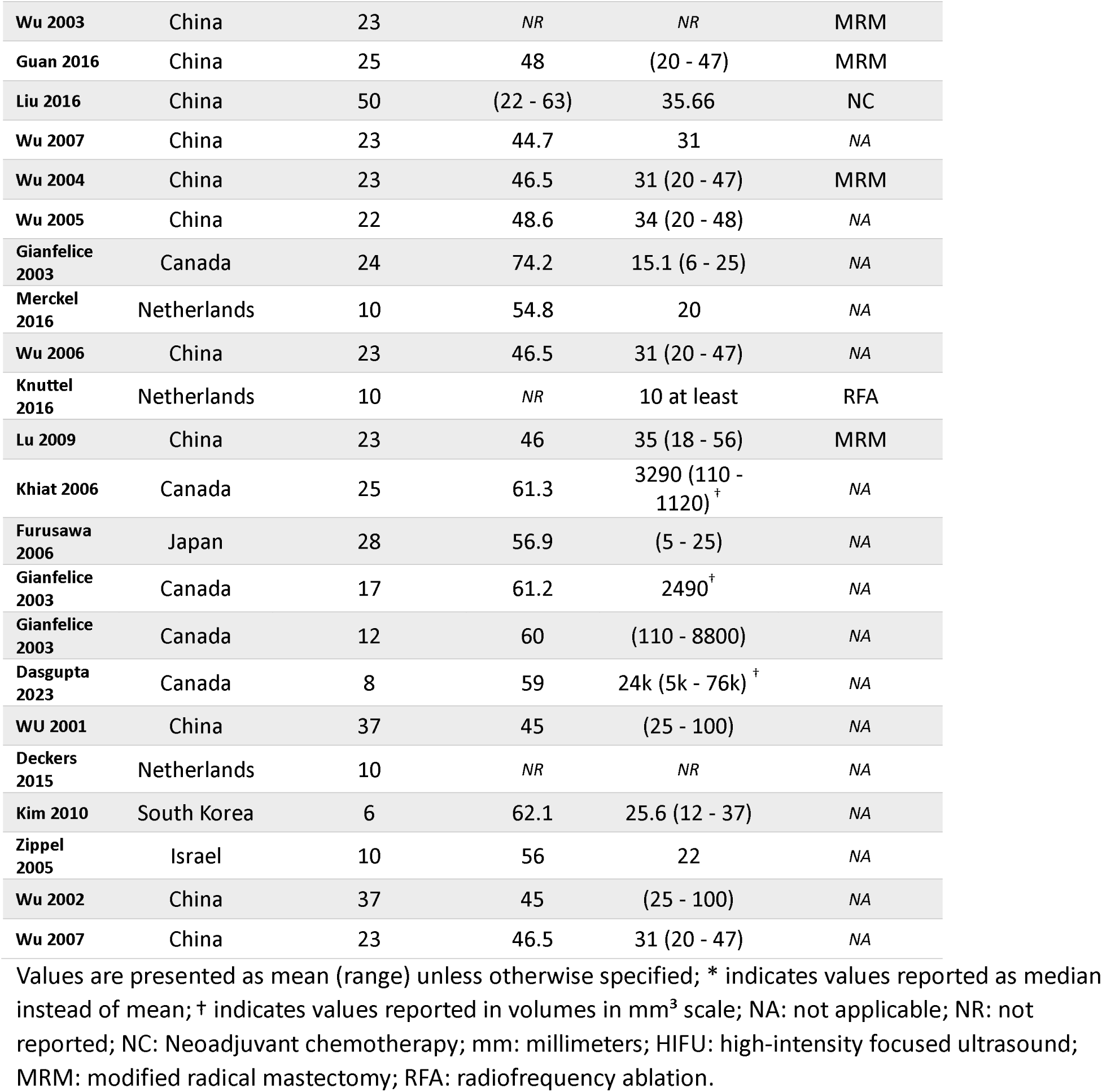
Study and patients’ characteristics of included studies.

### 3.3. Quality assessment

Nine randomized clinical trials (21–23,31,32,34,36,38,40) were assessed, and except one (38), all had some concerns regarding their risk of bias. The main domain in which all the studies except Guan et al. (38) had some concerns was the randomization process. However, the study by Wu et al. (22) also demonstrated some concerns regarding the outcome measurement (Figure S1).

Eighteen single-arm trials (16–20,23–30,33,35,37,39,41) were evaluated by the ROBINS-1 tool. Fourteen (16–19,24–30,33,37,41) were detected to have a low risk of bias. One study (39) had a moderate risk of bias due to some concerns in managing the missing data. The other three studies (17,20,32) did not provide sufficient information in managing the confounding factors; therefore, their risk of bias could not be estimated (Table S2).

### 3.4. Histopathological outcomes

Eighteen studies (16,18,19,21–30,33,35–38) reported histopathological results after the HIFU procedure based on the collected specimen. Four of them (21–23,29) investigated the expression of multiple biological markers such as CD44, proliferating cell nuclear antigen (PCNA), matrix metalloproteinase-9 (MMP-9), and heat-shock protein 70 (HSP-70). The results consistently showed that CD44, PCNA, and MMP-9 were not expressed in the HIFU group (21–23,29), whereas Wu et al. mentioned that HSP-70 positivity in the HIFU group was 100% (29). These studies reported similar positivity percentages of the markers in the control group (21–23,29). Other studies and Wu et al. (16,18,19,22,24–28,30,33,35–38) reported results on histopathological microscopic appearance of their specimen. Four studies (18,19,25,26) reported complete necrosis in less than 50% of their specimens. In contrast, five articles (22,27,28,30,37) found more than 50% complete necrosis in their specimens. Wu and colleagues (24) stated that coagulative necrosis appeared two weeks after the procedure. Additionally, they observed partial and complete fibrosis three months and 6-12 months after the treatment, respectively (24). Kim et al. (33) found no viable tumor in 50% of the specimens after 3-6 months, which increased to 67% during the next six months of their follow-up period. On the other hand, Liu et al. (35) studied the efficacy of HIFU as an add-on therapy to neoadjuvant chemotherapy in patients with breast cancer. Regarding the pathological analysis, they observed that HIFU improved the pathological complete response of neoadjuvant chemotherapy by 20.2% (35). Histopathological outcomes of the included studies are described in Table 2.

**Table 2.**
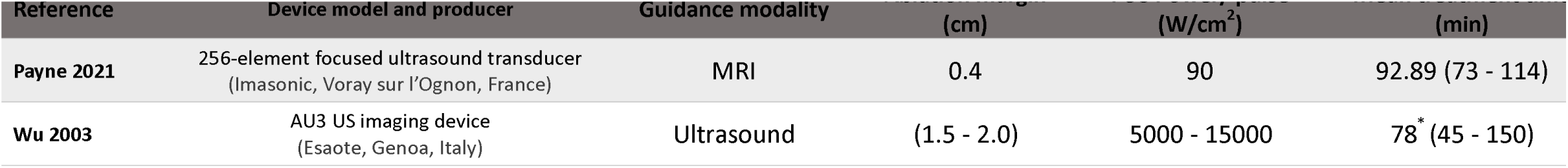
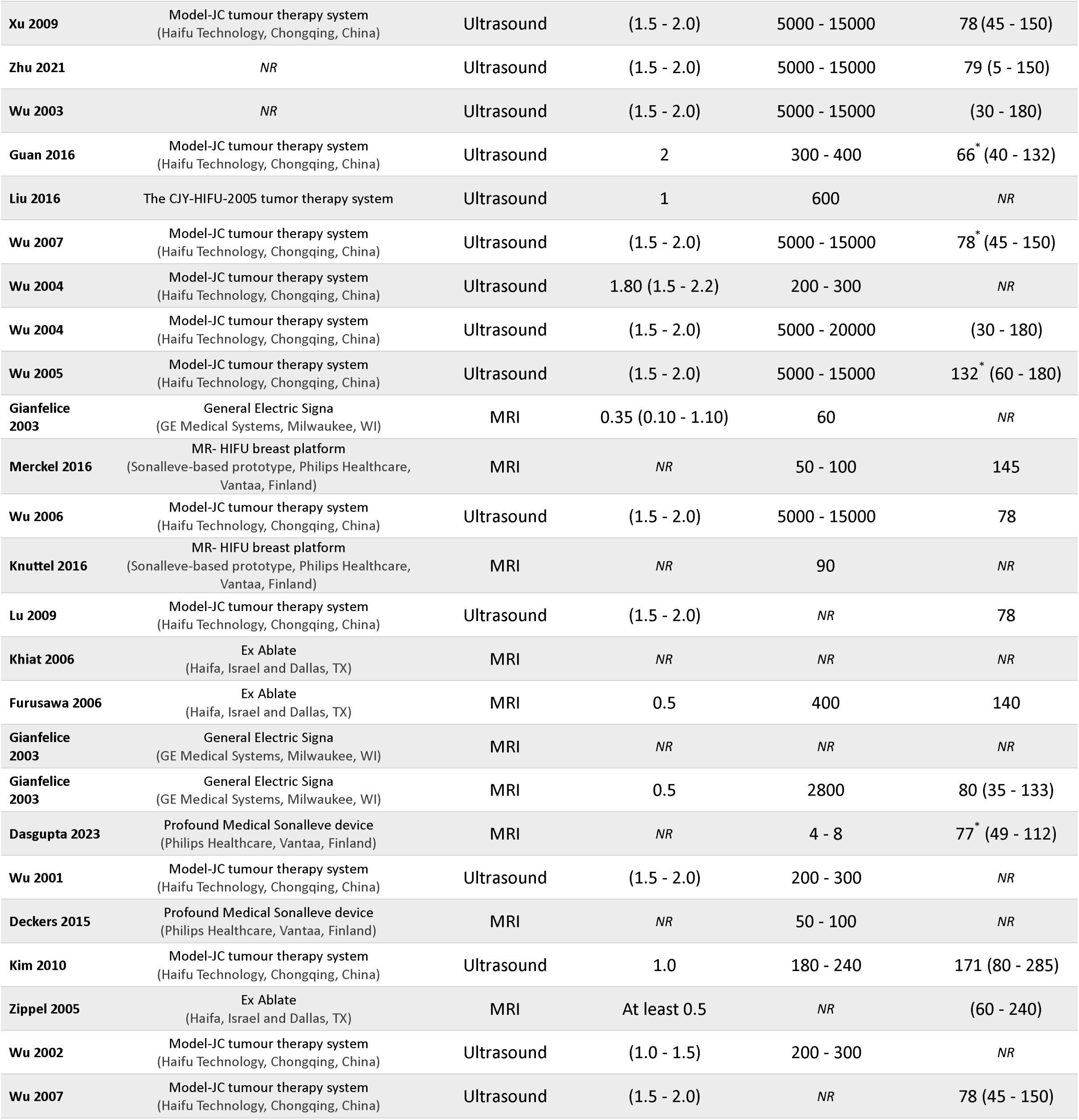
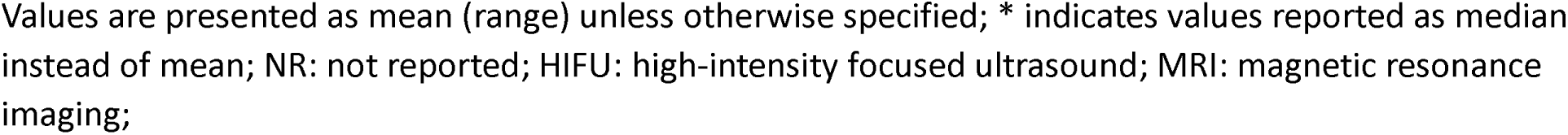
Treatment and HIFU device characteristics of included studies.

| Reference | Device model and producer | Guidance modality | Field size (cm) | Power (W/cm<sup>2</sup>) | Time (min) |
| --- | --- | --- | --- | --- | --- |
| Payne 2021 | 256-element focused ultrasound transducer (Imasonic, Voray sur l'Ognon, France) | MRI | 0.4 | 90 | 92.89 (73 - 114) |
| Wu 2003 | AU3 US imaging device (Esaote, Genoa, Italy) | Ultrasound | (1.5 - 2.0) | 5000 - 15000 | 78* (45 - 150) |

|  |  |  |  |  |  |
| --- | --- | --- | --- | --- | --- |
| **Xu 2009** | Model-JC tumour therapy system<br>(Haifu Technology, Chongqing, China) | Ultrasound | (1.5 - 2.0) | 5000 - 15000 | 78 (45 - 150) |
| **Zhu 2021** | *NR* | Ultrasound | (1.5 - 2.0) | 5000 - 15000 | 79 (5 - 150) |
| **Wu 2003** | *NR* | Ultrasound | (1.5 - 2.0) | 5000 - 15000 | (30 - 180) |
| **Guan 2016** | Model-JC tumour therapy system<br>(Haifu Technology, Chongqing, China) | Ultrasound | 2 | 300 - 400 | 66<sup>*</sup> (40 - 132) |
| **Liu 2016** | The CJY-HIFU-2005 tumor therapy system | Ultrasound | 1 | 600 | *NR* |
| **Wu 2007** | Model-JC tumour therapy system<br>(Haifu Technology, Chongqing, China) | Ultrasound | (1.5 - 2.0) | 5000 - 15000 | 78<sup>*</sup> (45 - 150) |
| **Wu 2004** | Model-JC tumour therapy system<br>(Haifu Technology, Chongqing, China) | Ultrasound | 1.80 (1.5 - 2.2) | 200 - 300 | *NR* |
| **Wu 2004** | Model-JC tumour therapy system<br>(Haifu Technology, Chongqing, China) | Ultrasound | (1.5 - 2.0) | 5000 - 20000 | (30 - 180) |
| **Wu 2005** | Model-JC tumour therapy system<br>(Haifu Technology, Chongqing, China) | Ultrasound | (1.5 - 2.0) | 5000 - 15000 | 132<sup>*</sup> (60 - 180) |
| **Gianfelice 2003** | General Electric Signa<br>(GE Medical Systems, Milwaukee, WI) | MRI | 0.35 (0.10 - 1.10) | 60 | *NR* |
| **Merckel 2016** | MR- HIFU breast platform<br>(Sonalleve-based prototype, Philips Healthcare,<br>Vantaa, Finland) | MRI | *NR* | 50 - 100 | 145 |
| **Wu 2006** | Model-JC tumour therapy system<br>(Haifu Technology, Chongqing, China) | Ultrasound | (1.5 - 2.0) | 5000 - 15000 | 78 |
| **Knuttel 2016** | MR- HIFU breast platform<br>(Sonalleve-based prototype, Philips Healthcare,<br>Vantaa, Finland) | MRI | *NR* | 90 | *NR* |
| **Lu 2009** | Model-JC tumour therapy system<br>(Haifu Technology, Chongqing, China) | Ultrasound | (1.5 - 2.0) | *NR* | 78 |
| **Khiat 2006** | Ex Ablate<br>(Haifa, Israel and Dallas, TX) | MRI | *NR* | *NR* | *NR* |
| **Furusawa 2006** | Ex Ablate<br>(Haifa, Israel and Dallas, TX) | MRI | 0.5 | 400 | 140 |
| **Gianfelice 2003** | General Electric Signa<br>(GE Medical Systems, Milwaukee, WI) | MRI | *NR* | *NR* | *NR* |
| **Gianfelice 2003** | General Electric Signa<br>(GE Medical Systems, Milwaukee, WI) | MRI | 0.5 | 2800 | 80 (35 - 133) |
| **Dasgupta 2023** | Profound Medical Sonalleve device<br>(Philips Healthcare, Vantaa, Finland) | MRI | *NR* | 4 - 8 | 77<sup>*</sup> (49 - 112) |
| **Wu 2001** | Model-JC tumour therapy system<br>(Haifu Technology, Chongqing, China) | Ultrasound | (1.5 - 2.0) | 200 - 300 | *NR* |
| **Deckers 2015** | Profound Medical Sonalleve device<br>(Philips Healthcare, Vantaa, Finland) | MRI | *NR* | 50 - 100 | *NR* |
| **Kim 2010** | Model-JC tumour therapy system<br>(Haifu Technology, Chongqing, China) | Ultrasound | 1.0 | 180 - 240 | 171 (80 - 285) |
| **Zippel 2005** | Ex Ablate<br>(Haifa, Israel and Dallas, TX) | MRI | At least 0.5 | *NR* | (60 - 240) |
| **Wu 2002** | Model-JC tumour therapy system<br>(Haifu Technology, Chongqing, China) | Ultrasound | (1.0 - 1.5) | 200 - 300 | *NR* |
| **Wu 2007** | Model-JC tumour therapy system<br>(Haifu Technology, Chongqing, China) | Ultrasound | (1.5 - 2.0) | *NR* | 78 (45 - 150) |

Values are presented as mean (range) unless otherwise specified; \* indicates values reported as median instead of mean; NR: not reported; HIFU: high-intensity focused ultrasound; MRI: magnetic resonance imaging;

### 3.5. MRI-based outcomes

Post-procedural MRI findings were mentioned in twelve studies (18,20,23,24,26,28,30,33,34,37,39,41). Eight studies (18,20,23,26,30,33,37,41) mainly discussed the difference in signal intensities or contrast enhancement in their study population. Of these, six studies (20,23,26,30,33,41) stated that no contrast enhancement was detected in the successfully treated regions. Furthermore, two (23,33) mentioned that a thin rim of enhancement implying necrosis or fibrosis in the tissue was observed. Likewise, four studies (18,20,26,33) showed that a persistent enhancement in MRI images correlates with a residual viable tumor. In the study conducted by Kim et al. (33), they observed a nodular irregular thick enhancement in partially ablated tumors. However, Merckel and colleagues (37) reported an incongruent result illustrating no differences in signal enhancement before and after the procedure. Payne et al. (39) performed a study evaluating the targeting algorithm and treatment planning for breast cancer using MR-guided focused ultrasound. They found that the average error between the desired and measured targeting in a phantom was 2.9±1.8, assessed either with MR-temperature imaging or MR-acoustic radiation force imaging. At the same time, 6.2±1.9 was achieved in previous studies. Similarly, Deckers et al. (34) investigated the targeting performance of MR-guided HIFU tumor ablation in breast cancer, demonstrating that the targeting accuracy and precision ranged from 2.4 to 2.6 mm and 1.5 to 1.8 mm, respectively.

### 3.6. Vessel destruction as an outcome for HIFU

Two studies (17,38) reported results on the effects of HIFU on the vasculature and blood supply of breast tumors. Wu et al. (17) stated that blood flow and microvasculature disturbance in the Color Doppler US were seen in ablated regions. Also, histological examination revealed a disruption in vascular elasticity and collagen fibrins (17). The latter was consistent with the findings of the study by Guan and colleagues (38), who also observed dispersed intravascular thrombin in the treated region. Moreover, they mentioned that the diameter of vessels was negatively correlated with HIFU-induced vascular damage (38).

### 3.7. Immunological responses

Three studies (31,32,40) stated outcomes related to immunological reactions to the HIFU procedure. Two of them showed that the ratio of CD4/CD8 cells was greater in the HIFU group (31,40). Lu et al. (31) observed that the numbers of T lymphocytes and tumor-infiltrating CD3, CD4, and CD8 were significantly greater in the ablated area in the HIFU group. Zhu and colleagues (40) investigated immune responses in tumor-draining lymph nodes and found significantly higher amounts of CD3, CD4, and CD57 cells in the HIFU group. In this study, they observed significantly different patterns of immune system reactivity in the HIFU group (40). Another study by Xu et l. (32) investigated the percentage of antigen-presenting cells (APC) after HIFU ablation. They noted a significant difference in the percentage of S-100, CD68, CD86, CD26, CD80, and HLA-DR between the two groups, demonstrating higher immune activity in the HIFU group (32).

### 3.8. Meta-analysis

From the included studies, 12 (18–20,24–26,28,33,35,37,39,41) provided sufficient information for meta-analysis of residual tumor proportion. Additionally, two studies (31,40) that reported immunological outcomes mentioned required data and entered a separate analysis.

As it is demonstrated in Figure 2, 223 patients were included in a random-effect analysis of the 12 studies. The weighted proportion of patients with residual tumor was 58.45 (95% CI; 45.48 – 71.42). However, the I^2^ statistics of 76% imply a possible substantial heterogeneity between the studies. On the other hand, we used a fixed-effect inverse-variance model to analyze mean differences in CD4/CD8 ratio between the HIFU and modified radical mastectomy group. The weighted mean difference of 0.6 (95% CI; 0.41 – 0.78) illustrates that CD4/CD8 ratio was significantly higher in the HIFU group. Moreover, the I^2^ statistics of 35.94 shows a moderate heterogeneity between the studies (Figure 3)

**Fig 2.**
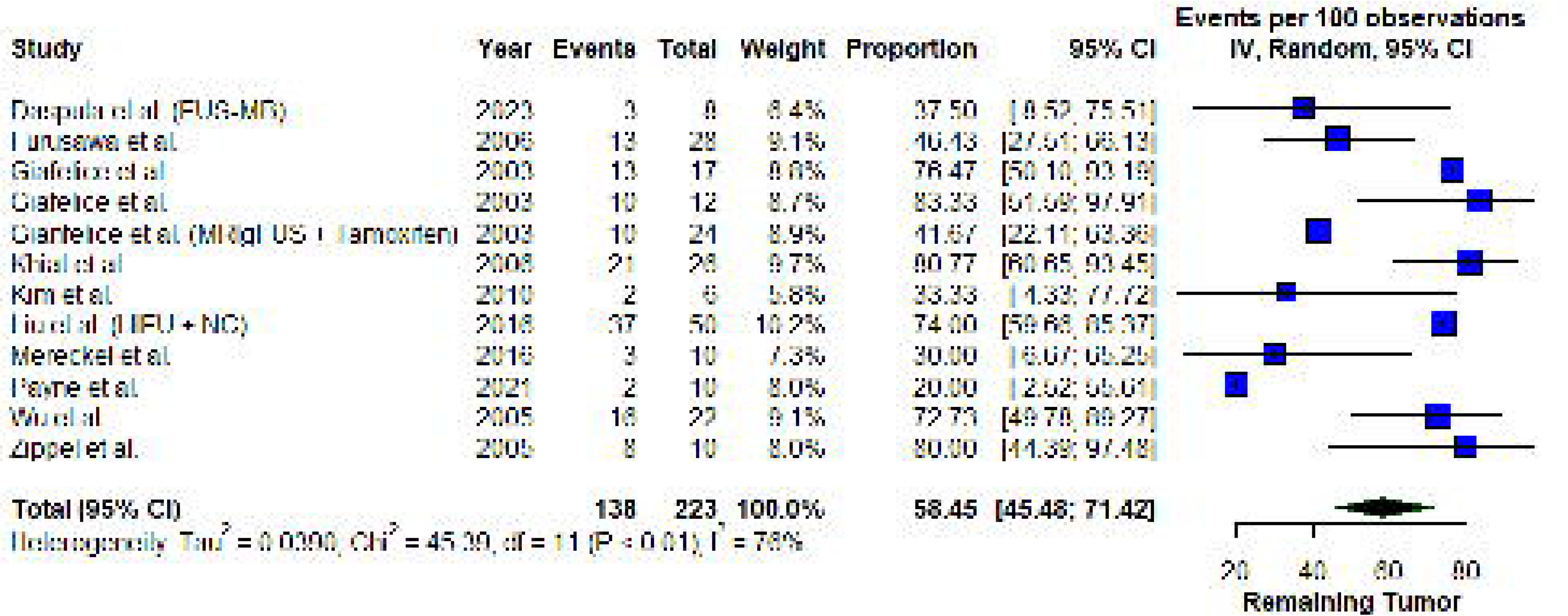
Forest Plot of Remaining Tumor Events Post-HIFU. This forest plot summarizes the proportion of remaining tumor events after treatment. Each study is listed with its publication year, the number of events, total sample size, weight, proportion of events, and 95% confidence interval (CI). The blue squares represent the effect size (proportion of remaining tumors) for each study, and the horizontal lines depict the 95% CI.

**Fig 3.**
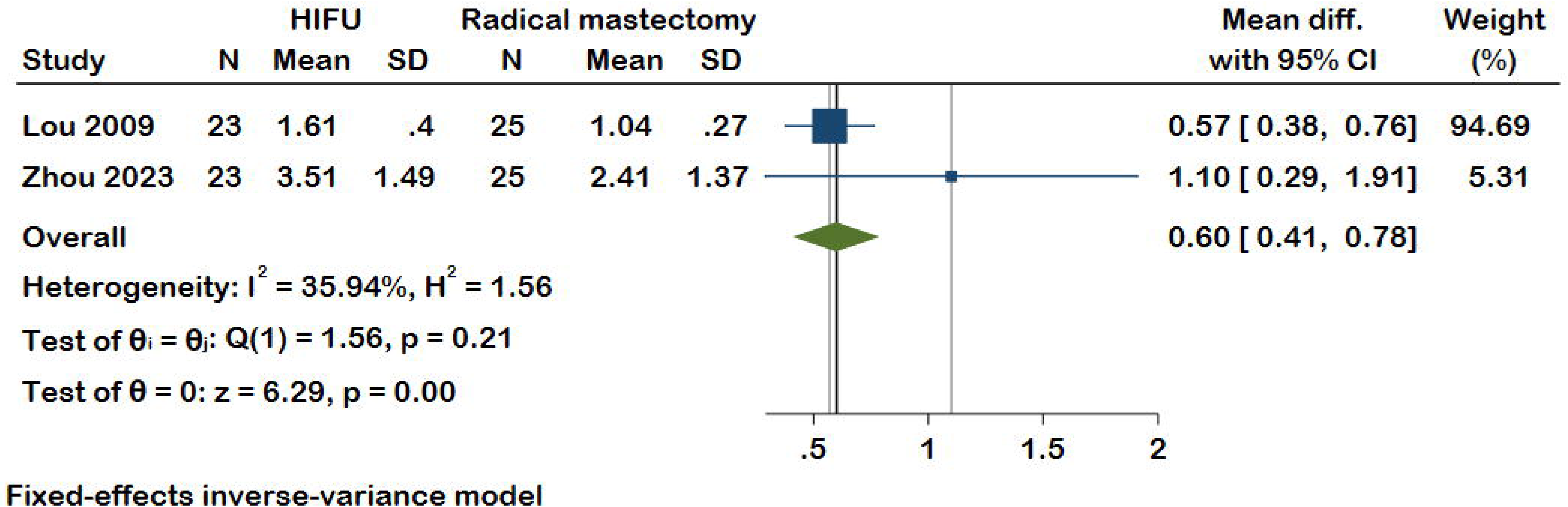
Forest Plot Comparing High-Intensity Focused Ultrasound (HIFU) and Radical Mastectomy in terms of CD4/CD8 ratio.

### 3.9. Follow-up imaging

Eight studies (20,22–24,26,33,38,41) reported information about the follow-up examination with vastly varied follow-up periods. Four studies (22,23,38,41) observed no contrast enhancement in the treated region, implying the absence of a viable tumor. However, Dasgupta et al. (41) found this observation in only three of 8 patients who underwent follow-up imaging. Also, in the study by Kim and colleagues (33), two of three patients in whom a complete ablation was achieved after the first HIFU session, no evidence of viable tumor was detected after 24-30 months of follow-up. At the same time, Gianfelice and colleagues (20) reported no significant changes in follow-up MRI images in 22 of 24 patients after one month.

Additionally, Wu et al. (23) performed a single-photon emission computed tomography (SPECT) scan in a short-term follow-up after one week, in which no radioisotope uptake was detected in the treated area. They also followed 22 patients for 1-2 years using Color and Power Doppler US, by which they observed a gradual shrinkage in the treated tissues. Furthermore, half of the patients showed total resorption of the tumor. In contrast, one patient presented with local recurrence after 18 months and underwent a modified radical mastectomy (23).

In another study in 2005, Wu and colleagues (24) followed their participants using US, MRI, and SPECT modalities. US images showed the disappearance of tumors in 8 patients and a reduction in size in 14 patients. In contrast, two patients presented local recurrence. Five patients underwent an MRI examination, and no contrast enhancement was observed. Moreover, no radioisotope uptake was detected in the SPECT images in the treated regions (24).

### 3.10. Side effects

Fifteen studies (19,20,22,24,25,27,28,30,33,34,36–39,41) reported data on HIFU-related complications. Figure 3 gives a better visual illustration of the prevalence of adverse effects of post-HIFU. Pain, as the most reported side effect, holds a percentage prevalence of 47.14%. The second major complication was skin burn, with a prevalence of 2.59%, although most burn cases were minimal or second-degree. However, one study (28) reported a third-degree skin burn in one patient. Furthermore, nausea and vomiting were the third most mentioned adverse effects related to the HIFU procedure. Other complications, such as cosmetic issues, fever, and edema, had a prevalence of less than 2%. Detailed information about side effects and their prevalence is shown in Table 1 and Figure 4.

**Fig 4.**
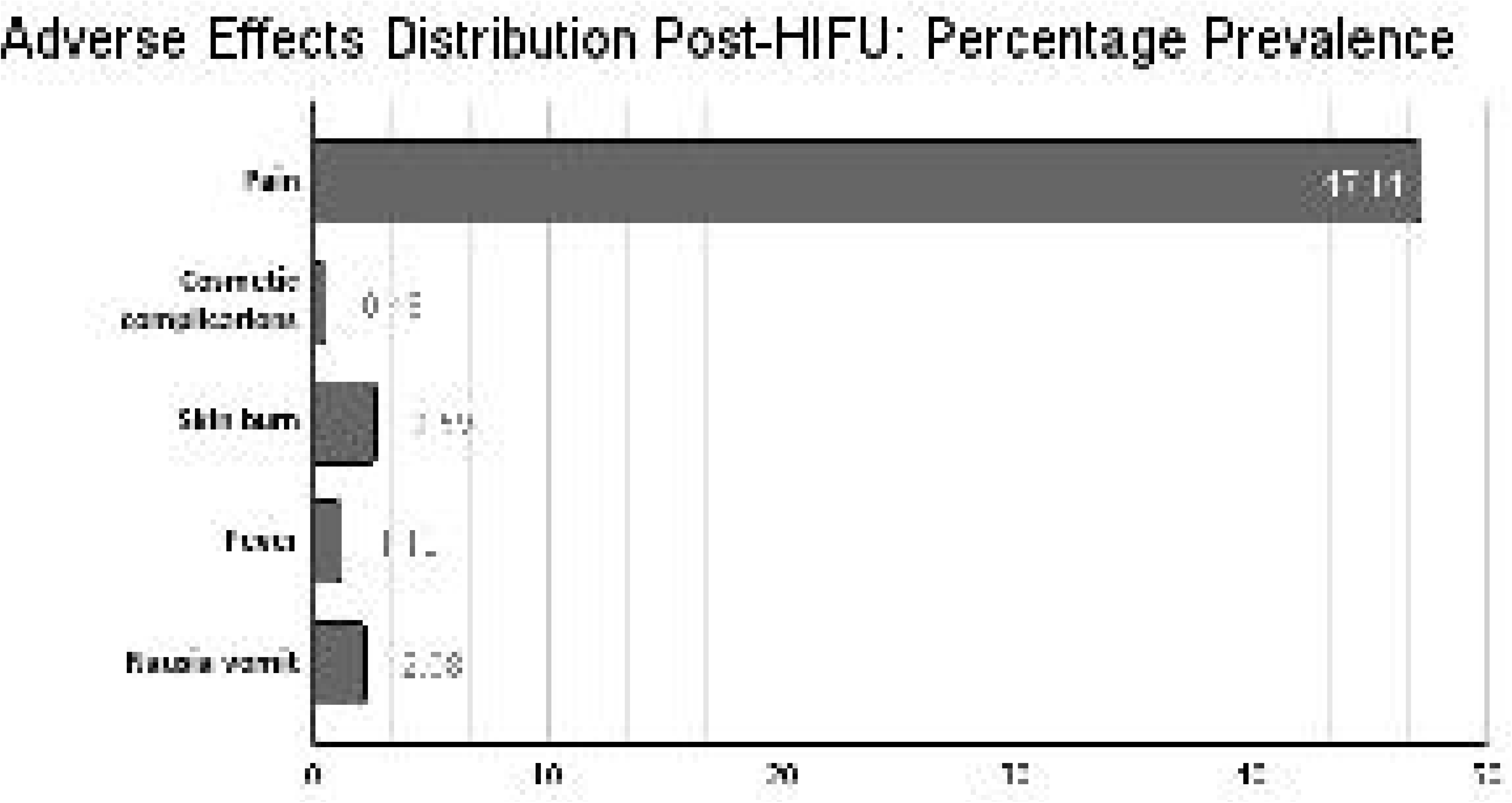
Adverse Effects Distribution Post-HIFU: Percentage Prevalence. The bar chart shows the percentage prevalence of various adverse effects observed after High-Intensity Focused Ultrasound (HIFU) treatment.

## 4. Discussion

In our systematic review, we analyzed 26 studies (16–41) involving 677 participants to investigate the impact of the HIFU procedure on malignant breast lesions. Our review included a meta-analysis of residual tumor proportion, reported in 12 studies (18–20,24–26,28,33,35,37,39,41), and a fixed-effect meta-analysis of CD4/CD8 ratio, reported in two studies (31,40). The results were categorized into four subheadings: histopathological, MRI-based, vessel destruction, and immunological outcomes. Key findings included the absence of CD44, MMP-9, and PCNA biological markers in specimens resected from the HIFU group (21–23,29) and a 100% positivity for HSP-70 expression (29). Most histopathological studies reported coagulative necrosis in the ablated area (18,19,22,25–28,30,37), and evidence suggests that HIFU ablation enhances the efficacy of neoadjuvant chemotherapy (35). MRI-based outcomes indicated changes in contrast enhancement correlated with the success of the HIFU procedure and a lower percentage of viable tumors (18,20,23,26,30,33,41). Studies also observed disturbances in blood flow and microvasculature of the targeted tissue due to vessel destruction by HIFU ablation (17,38). Furthermore, immunological responses indicated increased immune reactivity in the HIFU-ablated regions and an elevated CD4/CD8 ratio (31,32,40). Our meta-analysis showed a weighted proportion of 58.54% for the residual tumor proportion through random-effect analysis, with a relatively high heterogeneity.

Peek and colleagues (42) conducted a systematic review with objectives similar to ours, examining nine studies involving 167 patients. Their primary focus was on residual tumor percentage, as well as histopathological and imaging-based findings. They reported a 52.1% detection rate of residual tumor in their participants, which closely aligns with our findings of 58.54% residual tumor tissue. Additionally, Peek et al. (42) highlighted pain as the primary reported complication of the HIFU procedure, with a prevalence rate of 40.1%. They also observed a similar correlation between contrast enhancement and residual tumor. However, Peek et al. (42) did not incorporate some studies conducted before 2015, which are included in our review (16,17,22,23,27,29–32). An advantage of our study is that we explored the immunological responses to HIFU ablation and the procedure’s impact on vascular structure by including relevant studies. We also separately examined the short and long-term follow-up data presented in the studies (20,22–24,26,33,38,41).

In 2023, Zulkifli et al. (43) published a study that systematically reviewed research with similar objectives and reported almost identical findings concerning histopathological and imaging-based outcomes. However, they only included nine studies and, like Peek et al. (42), did not investigate the immunological and vascular outcomes of HIFU ablation. In addition, we addressed one of the issues raised in the study by Zulkifli and colleagues (43) by including two studies (34,39) that examined the efficacy of MR-guided HIFU ablation in breast cancer. Both studies demonstrated that the targeting accuracy of the MR-guided procedure was satisfactory (34,39). Furthermore, Payne et al. (39) proposed a targeting protocol and a treatment plan using MRTI and MR-ARFI as complementary tools for monitoring the treatment procedure. Another advantage of our study was the inclusion of patients with in situ carcinomas, a factor that was lacking in Zulkifli et al.’s study (43). Moreover, we set our exclusion threshold at studies with fewer than five patients, while Zulkifli and colleagues’ study (43) used a threshold of ten, resulting in the exclusion of five studies (33,34,36,39,41).

Previous studies have shown the essential role of CD4 T cells and CD4/CD8 ratio in immune reactivity against tumor tissue. Drescher et al. (44), in a review of tumor-infiltrating lymphocytes, stated that cytokine production is one of the critical functions of CD4 T cells, which induces the growth of CD8 cells. Besides, they mentioned that studies have proved that an increase in CD4 cell apoptosis is correlated with lower levels of TGF-β_1,_ which might be an evasion mechanism of malignant tumors (44). Likewise, Sheu and colleagues (45) enrolled 30 patients with cervical cancer and realized that lower CD4 levels and reversed CD4/CD8 ratio were consistent with rapid tumor growth and lymph node metastasis. On the other hand, Lu et al. (31) observed the infiltration of new T cells in the ablated area, leading to cytokine production and the development of cytotoxic T lymphocytes and NK cells. Similarly, Wu et al. (46) demonstrated some evidence in a review showing an increase in CD4/CD8 ratio one week after HIFU in choroidal melanoma and an increase in CD4 T cells, NK cells, and CD4/CD8 ratio after multiple session HIFU ablation in late-stage pancreatic cancer patients. Undoubtedly, our observations, showing a higher CD4/CD8 ratio in the HIFU group with a mean difference of 0.6, align with these findings and emphasize the importance of immunological responses following the HIFU procedure.

In our research, we conducted a random-effect meta-analysis to determine the weighted proportion of residual tumor and the extent of between-study heterogeneity, which was found to be relatively high. This high heterogeneity could be attributed to various factors related to the participants, interventions, and outcomes. Firstly, the participants in each study were not homogeneous, and different pathological types of breast cancer were included. Categorizing these types could potentially improve the heterogeneity. Secondly, a variety of interventions were implemented across the studies. Only one study (35) had a comparator group (neoadjuvant chemotherapy) and compared it with the combined therapy group (neoadjuvant chemotherapy + HIFU). Additionally, nine studies (18–20,25,26,28,37,39,41) utilized an MRI-guided procedure, while others (24,33,35) conducted US-guided HIFU ablation, leading to significant differences in the reported outcomes.

Our study had some limitations, which are discussed as follows: First, the included studies used different ablation methods, leading to heterogeneity between them. Second, the relatively small study population could potentially introduce bias in the trial outcomes. Third, the methods used in the included studies did not allow for assessing the independent effects of HIFU on tumors. Additionally, not all studies performed follow-up examinations to evaluate the survival rate of treated patients.

To conclude, the HIFU ablation procedure is considered a safe and effective treatment method for malignant breast cancer. It can be performed as an independent treatment or in conjunction with other methods. Its efficacy is attributed to its histopathological effects, immune reactivity, and vascular damage in the targeted area.

## Data Availability

Not applicable

## Figure legends

**Fig S1.**
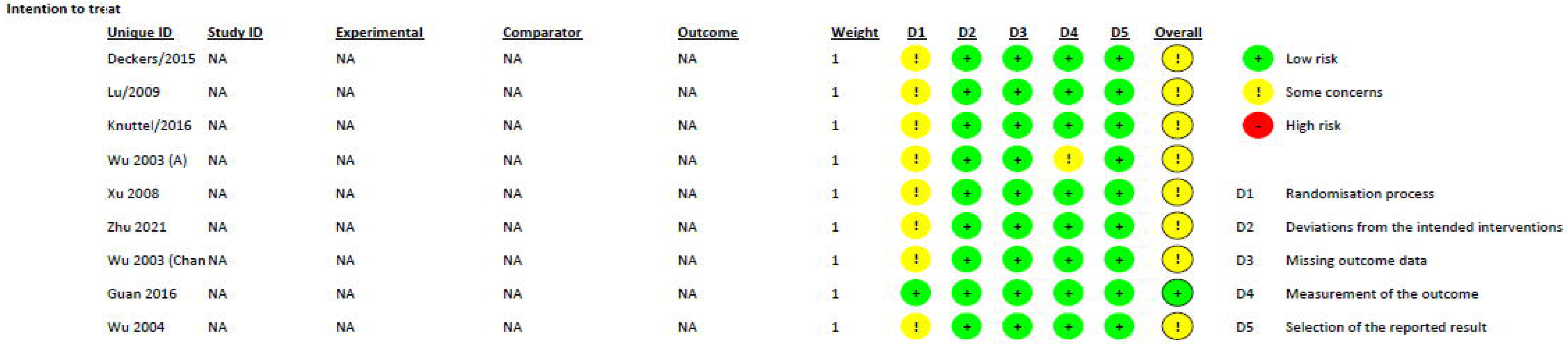
Risk of Bias Assessment for Randomized Clinical Trials. This table presents the results of a risk of bias assessment for various randomized clinical trials using five domains (D1-D5): D1 (randomization process), D2 (deviations from intended interventions), D3 (missing outcome data), D4 (measurement of the outcome), and D5 (selection of the reported result). Each domain is marked with a color-coded risk level: green (+) for low risk, yellow (!) for some concerns, and red (–) for high risk. The overall risk for each study is indicated in the "Overall" column.

