## Supplementary table 1 for "High-intensity focused ultrasound in treatment of primary breast cancer: a systematic review and meta-analysis"

**Table S1.** Search term for each database

**1. PubMed:** 477 (23 Dec 2023)

("Breast"[Mesh] OR "Breast Neoplasms"[Mesh] OR “breast”[Title/Abstract] OR "Breast Neoplasms"[Title/Abstract] OR "Breast Carcinoma In Situ"[Mesh] OR "Carcinoma, Ductal, Breast"[Mesh] OR "Carcinoma, Lobular"[Mesh] OR "Inflammatory Breast Neoplasms"[Mesh] OR "Triple Negative Breast Neoplasms"[Mesh] OR "Unilateral Breast Neoplasms"[Mesh]) AND ("High-Intensity Focused Ultrasound Ablation"[Mesh] OR "Ultrasonic Surgical Procedures"[Mesh] OR HIFU OR FUS OR "focused ultrasound ablation" OR “High-Intensity Focused Ultrasound” OR “focused US” OR “focused ultrasound” OR "Ultrasonic Surgical Procedures")

**2. Scopus:** 940 (23 Dec 2023)

TITLE-ABS-KEY(("Breast" OR "Breast Neoplasms" OR "breast" OR "Breast Neoplasms" OR "Breast Carcinoma In Situ" OR "Carcinoma, Ductal, Breast" OR "Carcinoma, Lobular" OR "Inflammatory Breast Neoplasms" OR "Triple Negative Breast Neoplasms" OR "Unilateral Breast Neoplasms" ) AND ("High-Intensity Focused Ultrasound Ablation" OR "Ultrasonic Surgical Procedures" OR HIFU OR FUS OR "focused ultrasound ablation" OR "High-Intensity Focused Ultrasound" OR "focused US" OR "focused ultrasound" OR "Ultrasonic Surgical Procedures"))

**3. Other sources (searching manually): 1**
