## Supplementary table 2 for "High-intensity focused ultrasound in treatment of primary breast cancer: a systematic review and meta-analysis"

Table S2. Quality Assessment of Included Studies Using the ROBINS-I Tool

| | confounding | selection of participants | classification of interventions | deviations from intended interventions | missing data | measurement of outcomes | selection of the reported result | Overall bias |
|---|---|---|---|---|---|---|---|---|
| Gianfelice/2003 TAMOXI | PY (1.1), N(1.2), NI (1.4, 1.6) | N(2.1), Y (2.4) | Y (3.1), Y (3.2), N (3.3) | N (4.1) | Y(5.1),N(5.2), N(5.3) | PN(6.1), NI(6.2),Y(6.3), N(6.4) | N,N,N | No information |
| Merckel/2016 | PN | N(2.1), Y (2.4) | Y (3.1), Y (3.2), N (3.3) | N (4.1) | Y(5.1),N(5.2), N(5.3) | PN(6.1), NI(6.2),Y(6.3), N(6.4) | N,N,N | LOW-RISK OF BIAS |
| Feng Wu/2006 heat fix | PN | N(2.1), Y (2.4) | Y (3.1), Y (3.2), N (3.3) | N (4.1) | Y(5.1),N(5.2), N(5.3) | PN(6.1), NI6.2),Y(6.3), N(6.4) | N,N,N | LOW-RISK OF BIAS |
| KHIAT/2006 | PN | N(2.1), Y (2.4) | Y (3.1), Y (3.2), N (3.3) | N (4.1) | Y(5.1),N(5.2), N(5.3) | PN(6.1), NI(6.2),Y(6.3), N(6.4) | N,N,N | LOW-RISK OF BIAS |
| Furusawa/2006 | PN | N(2.1), Y (2.4) | Y (3.1), Y (3.2), N (3.3) | N (4.1) | Y(5.1),N(5.2), N(5.3) | PN(6.1), NI(6.2),Y(6.3), N(6.4) | N,N,N | LOW-RISK OF BIAS |
| Gianfelice / 2003 | PN | N(2.1), Y (2.4) | Y (3.1), Y (3.2), N (3.3) | N (4.1) | Y(5.1),N(5.2), N(5.3) | PN(6.1), NI(6.2),Y(6.3), N(6.4) | N,N,N | LOW-RISK OF BIAS |
| Gianfelice / 2003 Effectiveness | PN | N(2.1), Y (2.4) | Y (3.1), Y (3.2), N (3.3) | N (4.1) | Y(5.1),N(5.2), N(5.3) | PN(6.1), NI(6.2),Y(6.3), N(6.4) | N,N,N | LOW-RISK OF BIAS |
| Dasgupta/2023 | PN | N(2.1), Y (2.4) | Y (3.1), Y (3.2), N (3.3) | N (4.1) | Y(5.1),N(5.2), N(5.3) | PN(6.1), NI(6.2),Y(6.3), N(6.4) | N,N,N | LOW-RISK OF BIAS |
| WU/2001/ pathological change | PN | N(2.1), Y (2.4) | Y (3.1), Y (3.2), N (3.3) | N (4.1) | Y(5.1),N(5.2), N(5.3) | PN(6.1), NI(6.2),Y(6.3), N(6.4) | N,N,N | LOW-RISK OF BIAS |
| KIM/2006 | PN | N(2.1), Y (2.4) | Y (3.1), Y (3.2), N (3.3) | N (4.1) | Y(5.1),N(5.2), N(5.3) | PN(6.1), NI(6.2),Y(6.3), N(6.4) | N,N,N | LOW-RISK OF BIAS |
| Zippel B/2005 | PN | N(2.1), Y (2.4) | Y (3.1), Y (3.2), N (3.3) | N (4.1) | Y(5.1),N(5.2), N(5.3) | PN(6.1), NI(6.2),Y(6.3), N(6.4) | N,N,N | LOW-RISK OF BIAS |
| Feng Wu/2002 TUMOR VESSEL DESTRUCTION | PN | N(2.1), Y (2.4) | Y (3.1), Y (3.2), N (3.3) | N (4.1) | Y(5.1),N(5.2), N(5.3) | PN(6.1), NI(6.2),Y(6.3), N(6.4) | N,N,N | LOW-RISK OF BIAS |
| FENG WU / 2007/ Wide Local Ablation | PN | N(2.1), Y (2.4) | Y (3.1), Y (3.2), N (3.3) | N (4.1) | Y(5.1),N(5.2), N(5.3) | PN(6.1), NI(6.2),Y(6.3), N(6.4) | N,N,N | LOW-RISK OF BIAS |
| Payne/2021 | PN | N(2.1), Y (2.4) | Y (3.1), Y (3.2), N (3.3) | N (4.1) | PN(5.1),Y(5.2) ,N(5.3),Y(5.4), PY(5.5) | N(6.1), Y(6.2),Y(6.3), N(6.4) | N,N,N | MODERAT-RISK OF BIAS |
| Liu 2016 | PY (1.1), N(1.2), NI (1.4, 1.6) | N(2.1), Y (2.4) | Y (3.1), Y (3.2), N (3.3) | N (4.1) | Y(5.1),N(5.2), N(5.3) | PN(6.1), NI(6.2),Y(6.3), N(6.4) | N,N,N | No information |
| Wu 2007 | N | N(2.1), Y (2.4) | Y (3.1), Y (3.2), N (3.3) | N (4.1) | Y(5.1),N(5.2), N(5.3) | N(6.1), Y(6.2),Y(6.3), N(6.4) | N,N,N | LOW-RISK OF BIAS |
| Wu 2004 | PY (1.1), N(1.2), NI (1.4, 1.6) | N(2.1), Y (2.4) | Y (3.1), PY (3.2), PN (3.3) | N (4.1) | Y(5.1),PN(5.2) ,PN(5.3) | N(6.1), Y(6.2),Y(6.3), N(6.4) | N,N,N | No information |
| Wu 2004 (overview) | PY (1.1), N(1.2), NI (1.4, 1.6) | N(2.1), Y (2.4) | Y (3.1), PY (3.2), PN (3.3) | N (4.1) | Y(5.1),PN(5.2) ,PN(5.3) | N(6.1), Y(6.2),Y(6.3), N(6.4) | N,N,N | No information |

| | confounding | selection of participants | classification of interventions | deviations from intended interventions | missing data | measurement of outcomes | selection of the reported result | Overall bias |
|---|---|---|---|---|---|---|---|---|
| Wu 2005 | PN | **N**(2.1), **Y** (2.4) | **Y** (3.1), **Y** (3.2), N (3.3) | Y (4.1), N (4.2) | PY(5.1),PN(5.2),PN(5.3) | PN(6.1), Y(6.2),Y(6.3), N(6.4) | N,N,N | LOW-RISK OF BIAS |

.Abbreviations: PN, Probably No; PY, Probably Yes; N, No; Y, Yes; NI, No Information
